# Prevalence and factors associated with hepatitis B surface antibody seroprotection among healthcare workers undergoing laboratory testing at Eastgate Clinic, Windhoek, Namibia: A retrospective cross-sectional study

**DOI:** 10.64898/2026.10.01.26364556

**Authors:** Talent Farai Mafumhe, Vonai Charamba, Onyemaechi Okpara Azu, Hamufare Dumisani Mugauri

**Author notes:** Corresponding author: Hamufare Dumisani Mugauri, Department of Global Public Health and Family Medicine, Faculty of Medicine and Health Sciences, University of Zimbabwe, Harare, Zimbabwe.

## Abstract

Healthcare workers are occupationally exposed to blood and body fluids and require effective protection against hepatitis B virus infection. Evidence on hepatitis B surface antibody (anti-HBs) seroprotection among healthcare workers in Namibia remains limited. We estimated the prevalence of anti-HBs seroprotection and examined associated factors among healthcare workers tested at Eastgate Clinic in Windhoek, Namibia. We conducted a cross-sectional analysis of routinely collected anti-HBs test records from 1 January to 31 December 2025. The first chronological test per individual was included. Anti-HBs concentrations ≥10 mIU/mL were classified as seroprotective. Results outside the assay reporting range were treated as censored but remained classifiable for the binary outcome. Prevalence estimates were reported with Wilson 95% confidence intervals (CIs). Modified Poisson regression with robust standard errors was used to estimate adjusted prevalence ratios (aPRs). The register contained 304 test records representing 299 unique individuals. After excluding three individuals with missing anti-HBs results, 296 were analysed; 163 (55.1%) were female. Overall, 148 individuals were seroprotected, corresponding to a prevalence of 50.0% (95% CI 44.3–55.7%). Seroprotection was lowest among participants aged 18– 29 years (35.3%) and highest among those aged 50–59 years (71.1%). Each 10-year increase in age was associated with a higher prevalence of seroprotection after adjustment for sex (aPR 1.14, 95% CI 1.06–1.24; p=0.001), whereas sex was not independently associated with seroprotection (aPR 0.97, 95% CI 0.78–1.22; p=0.797). The age association remained consistent across sensitivity analyses. Half of the healthcare workers tested had anti-HBs concentrations below the operational seroprotection threshold. These findings support systematic vaccination verification, appropriately timed post-vaccination testing, and documented follow-up of non-protective results. The absence of vaccination history, occupational characteristics, testing indications, and additional hepatitis B serological markers limited causal and clinical interpretation.

## Introduction

Hepatitis B virus (HBV) infection remains a major cause of acute hepatitis, chronic liver disease, cirrhosis, and hepatocellular carcinoma worldwide. Vaccination is the principal preventive intervention, with global policy emphasising universal infant immunisation and vaccination of populations at increased risk of occupational or other exposure [1,2]. Healthcare workers constitute an important risk group because their clinical and laboratory duties may involve percutaneous, mucosal, or non-intact skin exposure to blood and other potentially infectious body fluids [3,4]. Ensuring adequate hepatitis B protection among healthcare workers is therefore integral to occupational health, infection prevention and control, and the continuity and safety of healthcare delivery [2].

The occupational burden of HBV is particularly relevant in sub-Saharan Africa, where the background prevalence of infection remains comparatively high and healthcare workers may encounter persistent barriers to vaccination, post-vaccination testing, and post-exposure management [3–6]. Studies across Africa have documented substantial variation in hepatitis B vaccination uptake, incomplete vaccination schedules, and uneven awareness of occupational risk and prevention practices among healthcare workers [4–6]. Institutional constraints, including limited access to vaccines, inadequate documentation of vaccination status, and inconsistent implementation of occupational-health policies, may further compromise protection [4,6]. In Namibia, previous research found that healthcare workers were generally aware of hepatitis B, but that the availability and implementation of national guidance for prevention and control were insufficient [7]. Despite this recognised occupational risk, empirical evidence on hepatitis B serological protection among healthcare workers in Namibia remains limited.

Antibody to hepatitis B surface antigen (anti-HBs) is commonly measured to assess serological response following hepatitis B vaccination. An anti-HBs concentration of at least 10 mIU/mL, measured at an appropriate interval after completion of a documented vaccination series, is conventionally considered evidence of an adequate serological response [1,8]. However, interpretation of anti-HBs outside a documented post-vaccination context requires caution. Antibody concentrations may decline over time, and a low or undetectable anti-HBs concentration does not necessarily indicate loss of immune memory in an immunocompetent person who previously achieved an adequate response. Conversely, a positive anti-HBs result alone cannot distinguish vaccine-induced immunity from immunity following resolved natural infection. Additional markers, particularly hepatitis B surface antigen (HBsAg) and total antibody to hepatitis B core antigen (anti-HBc), are required to characterise HBV infection and immunity more comprehensively [8,9].

Routine laboratory records can provide useful evidence on the distribution of anti-HBs concentrations among individuals who undergo testing, especially in settings where integrated occupational-health surveillance systems are not available. Such data can identify groups with antibody concentrations below the operational seroprotection threshold and inform the organisation of vaccination verification, post-vaccination testing, and clinical follow-up. However, laboratory registers describe a selected testing population rather than a probability sample of the broader healthcare workforce. Estimates derived from these records must therefore be interpreted as the prevalence of seroprotection among healthcare workers undergoing testing, rather than as workforce-wide vaccination coverage or immunity prevalence.

This study estimated the prevalence of anti-HBs seroprotection among healthcare workers undergoing laboratory testing at Eastgate Clinic in Windhoek, Namibia, during 2025. It also assessed whether seroprotection was associated with age, sex, and calendar period. We defined seroprotection operationally as an anti-HBs concentration of at least 10 mIU/mL and examined the robustness of the observed associations across alternative analytical specifications.

## Materials and methods

### Study design and setting

We conducted a retrospective laboratory-based cross-sectional study using routinely collected hepatitis B surface antibody (anti-HBs) test records from Eastgate Clinic in Windhoek, Namibia. Eastgate Clinic is a private healthcare facility providing clinical, laboratory, and pharmacy services. The study covered anti-HBs tests performed between 1 January and 31 December 2025. The study was reported in accordance with the Strengthening the Reporting of Observational Studies in Epidemiology statement and the RECORD extension for observational studies conducted using routinely collected health data [11,12].

### Study population and eligibility criteria

The source population comprised healthcare workers who underwent quantitative anti-HBs testing at Eastgate Clinic during the study period. Healthcare-worker status was verified against the occupation and referral information recorded on the original laboratory request forms. Records were included only when the request documentation identified the individual as a healthcare worker, healthcare trainee, or other staff member working in a healthcare setting. Records for which healthcare-worker status could not be verified were excluded.

Records without a reported anti-HBs result were retained for the participant-flow description but excluded from analyses requiring seroprotection status. Where more than one test was identified for the same individual, the first chronological test was included in the primary cross-sectional analysis to preserve the independence of observations. Subsequent test records were retained for record-linkage and data-quality assessment but were not analysed as independent observations.

### Data sources and variable definitions

Data were accessed for research purposes on 14 September 2026 and extracted from the routine laboratory register into a structured dataset. The source variables were test date, laboratory accession number, patient name, sex, age, and anti-HBs result. Authorised members of the study team had temporary access to identifiable participant information, including names and laboratory accession numbers, during data extraction, source verification, identification of repeat tests, and reconciliation with source documents. Each eligible individual was subsequently assigned a unique study identifier, after which names and other direct identifiers were removed from the analytical dataset. The identifiable linkage file was stored separately under restricted access and was not used in the statistical analyses. Only the de-identified analytical dataset was available during analysis and preparation of the study findings.

Age was analysed as a continuous variable and was also categorised as <18, 18–29, 30–39, 40–49, 50–59, and ≥60 years for descriptive analysis. Sex was analysed as recorded in the laboratory register. Calendar month and quarter were derived from the verified test date.

The routine laboratory dataset did not contain healthcare-worker cadre, employing facility, degree of patient contact, duration of healthcare employment, indication for anti-HBs testing, hepatitis B vaccination history, number or dates of vaccine doses, time since the final vaccine dose, or whether testing was performed before or after vaccination. Information on HIV status, immunosuppressive conditions, hepatitis B surface antigen (HBsAg), total antibody to hepatitis B core antigen (anti-HBc), and clinical management following a non-protective result was also unavailable. These characteristics were consequently not included in the statistical models.

### Laboratory testing and outcome definition

Anti-HBs concentrations were measured using the Elecsys Anti-HBs II electrochemiluminescence immunoassay on a cobas e 411 immunoassay analyser (Roche Diagnostics GmbH, Mannheim, Germany), using serum or plasma specimens. Results were reported in milli-international units per millilitre. The analytical measurement range was 2–1000 mIU/mL; concentrations below and above this range were reported as <2 mIU/mL and >1000 mIU/mL, respectively. Testing was performed in accordance with the manufacturer’s instructions and Eastgate Clinic laboratory standard operating procedures. Internal quality control was performed at the beginning of each analytical run and following reagent-lot changes, calibration, or instrument maintenance, using negative and positive anti-HBs control materials. Calibration was performed using manufacturer-supplied calibration material following reagent-lot changes, when prompted by the analyser, and when quality-control results fell outside predefined acceptance limits. The analyser was maintained in accordance with the manufacturer’s preventive-maintenance schedule.

The primary outcome was anti-HBs seroprotection, defined operationally as a concentration ≥10 mIU/mL. Results <10 mIU/mL were classified as non-protective, while results ≥10 mIU/mL were classified as seroprotective. Values reported as <2 mIU/mL were therefore classified as non-protective, and values reported as >1000 mIU/mL were classified as seroprotective. Because these observations were censored at the assay reporting limits, they were not treated as exact concentrations in analyses of the quantitative distribution.

For descriptive purposes, anti-HBs results were grouped as <2, 2–9.9, 10–99.9, 100–1000, and >1000 mIU/mL. The study did not interpret anti-HBs positivity as evidence of vaccine-derived immunity because vaccination history, HBsAg, and total anti-HBc results were unavailable.

### Data cleaning and management

The extracted data were assessed for completeness, internal consistency, duplicate records, implausible values, and chronological inconsistencies. Blank spreadsheet rows were removed, sex entries were standardised, age was converted to completed years, and anti-HBs results were parsed while preserving their original reporting format.

Potential repeat tests were identified by comparing normalised names, age, sex, and chronological laboratory records. Five individuals had two apparent test records. The first chronological test was retained for the primary analysis, while all records were included in a sensitivity analysis. One apparent repeat pair contained discordant sex entries and was flagged for verification against the source register rather than being corrected by assumption.

Five test dates were inconsistent with the 2025 study period or the chronological sequence of laboratory accession numbers. Plausible dates were provisionally assigned using the dates and accession numbers of adjacent laboratory records, and the affected observations were flagged in the data-cleaning log for source verification. These provisional corrections did not affect classification of the primary outcome or the principal age-and-sex regression model because seroprotection status was determined from the anti-HBs result and test date was not included in the primary model. Analyses involving calendar quarter should therefore be interpreted with appropriate caution.

The de-identified analytical dataset was stored separately from the linkage file containing names and laboratory accession numbers. The linkage file was accessible only to authorised study personnel and was not used in statistical modelling.

### Statistical analysis

Participant characteristics were summarised using frequencies and percentages for categorical variables. Age was summarised using the mean and standard deviation and the median and interquartile range. Anti-HBs results were described using censoring-aware categories because measurements reported below or above the assay range did not represent exact concentrations.

The prevalence of seroprotection was calculated as the number of individuals with anti-HBs ≥10 mIU/mL divided by the number with an evaluable anti-HBs result. Prevalence estimates were reported with Wilson 95% confidence intervals (CIs). Seroprotection was described overall and according to sex, age group, and calendar quarter.

Because seroprotection was a common outcome, associations were estimated using modified Poisson regression with robust standard errors and reported as prevalence ratios (PRs) with 95% CIs. The primary multivariable model included age and sex. Age was modelled continuously per 10-year increase to preserve information, avoid arbitrary categorisation, and limit the number of model parameters. Sex was entered as a binary categorical variable, with male participants as the reference category.

Calendar quarter was added to the primary model to assess whether temporal variation in the composition of individuals tested materially altered the age and sex estimates. A second sensitivity analysis restricted the analytical population to participants aged ≥18 years. Age was also modelled categorically, using participants aged 18–29 years as the reference group, to assess the robustness of the primary continuous-age specification. Departure from linearity was evaluated by adding a quadratic age term to the primary model. Repeat test records were not included as independent observations because doing so would violate the independence assumption.

No automated or statistical-significance-based variable-selection procedures were used. Age and sex were included a priori because they were the only individual-level explanatory variables available consistently in the routine dataset.

Model estimates were exponentiated and reported as PRs or adjusted PRs with 95% CIs. Statistical tests were two-sided, and p<0.05 was considered statistically significant. No automated variable-selection procedures were used. Analyses were conducted using Python version 3.12.14, pandas version 2.2.2, SciPy version 1.13.1, and statsmodels version 0.15.0.

### Ethical considerations

The Eastgate Clinic Institutional Review Board reviewed the study and determined that it qualified for exemption from full ethical review because it involved retrospective secondary analysis of routinely collected laboratory records, no participant recruitment or direct contact, no intervention, and no alteration of clinical care. The Board waived the requirement for individual informed consent because retrospective recontact was impracticable and the study presented no more than minimal risk. The exemption and waiver were granted on 28 September 2026 under reference number 246/2025. Eastgate Clinic authorised the use of the de-identified records and publication of aggregate findings.

The analytical dataset was de-identified before analysis. Direct identifiers, including names and laboratory accession numbers, were retained only in a separate, access-restricted linkage file used for source verification and identification of repeat records. Results were reported in aggregate, and no individual healthcare worker was identifiable in the manuscript or supporting materials. The principal foreseeable risk was breach of confidentiality; this risk was mitigated through data minimisation, de-identification, restricted access, secure storage, and aggregation of potentially identifiable subgroups.

## Results

### Participant selection and analytical population

The source register contained 304 anti-HBs test records. Linkage using normalised personal identifiers identified 299 unique individuals, five of whom had two test records. For these individuals, the first chronological test was retained in the primary cross-sectional analysis. Three individuals had no reported anti-HBs result and were excluded from analyses requiring seroprotection status. The primary complete-case analysis therefore included 296 participants. Eight participants were younger than 18 years, leaving 288 participants in the adult-only sensitivity analysis **(Fig 1).**

**Fig 1.**
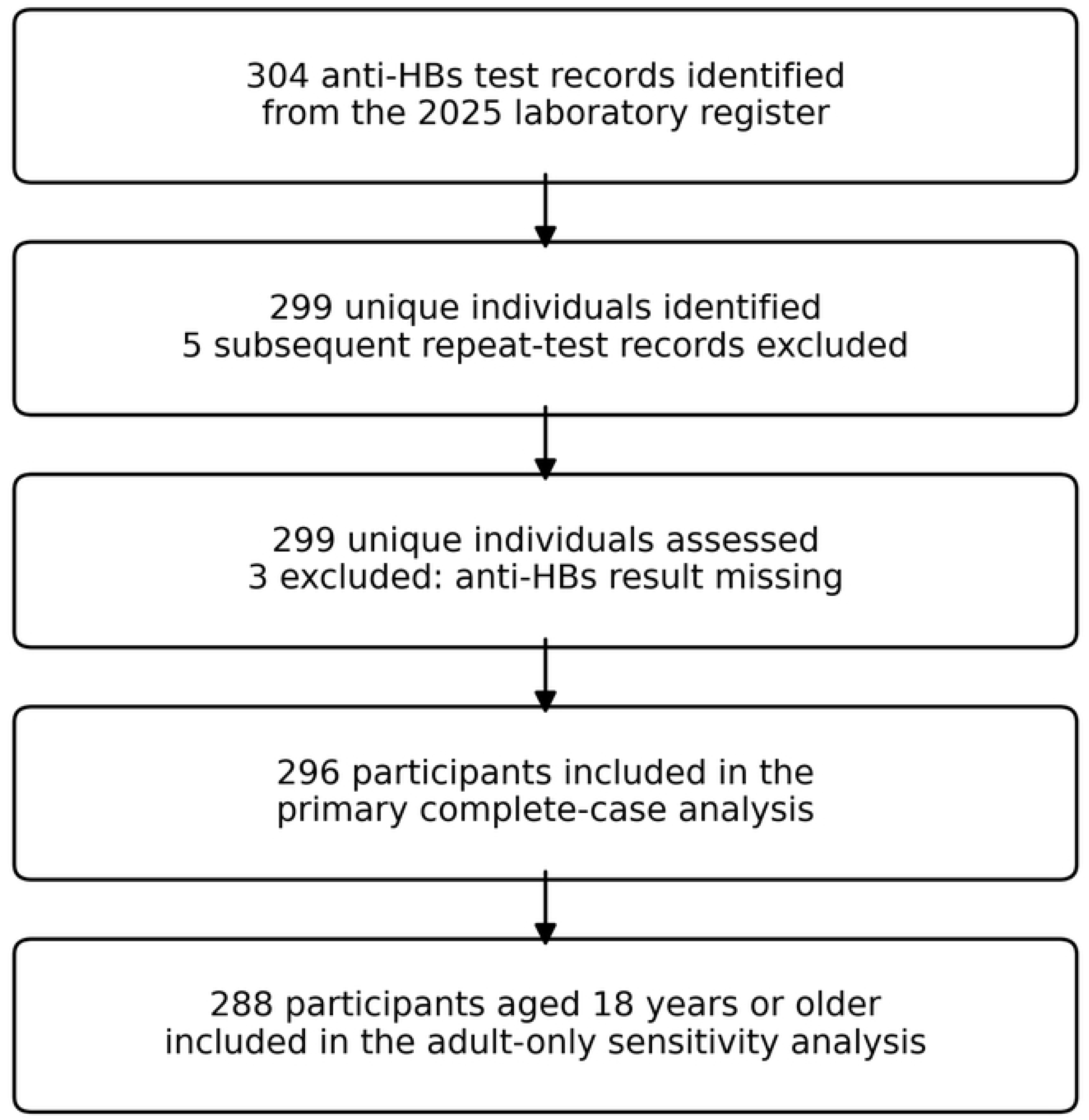
Selection of participants for the primary and sensitivity analyses. The source register contained 304 anti-HBs test records representing 299 unique individuals. The first chronological test was retained for each individual. Three individuals without a reported anti-HBs result were excluded from outcome analyses, leaving 296 participants in the primary complete-case analysis. Of these, 288 were aged at least 18 years and were included in the adult-only sensitivity analysis. Anti-HBs, antibody to hepatitis B surface antigen.

The median age of participants was 36 years (interquartile range [IQR] 29–49; range 14–81), and the mean age was 38.9 years (standard deviation 13.4). Of the 296 participants, 163 (55.1%) were female, and 133 (44.9%) were male. Participants with anti-HBs concentrations below 10 mIU/mL had a median age of 34 years (IQR 27–44), compared with 41 years (IQR 32–51) among participants with seroprotective concentrations.

### Distribution of anti-HBs results

Among the 296 participants in the primary analysis, 132 (44.6%) had anti-HBs results below the lower reporting limit of 2 mIU/mL, and 16 (5.4%) had concentrations between 2 and 9.9 mIU/mL. Among participants meeting the seroprotection threshold, 40 (13.5%) had concentrations of 10–99.9 mIU/mL, 72 (24.3%) had concentrations of 100–1000 mIU/mL, and 36 (12.2%) had results above the upper reporting limit of 1000 mIU/mL. Thus, 148 participants had anti-HBs concentrations below 10 mIU/mL and 148 had concentrations ≥10 mIU/mL.

Across all 304 laboratory records, including repeat tests and records with missing outcomes, 133 results were reported below the lower reporting limit, 37 above the upper reporting limit, 131 as exact quantitative measurements, and three were missing. Consequently, 170 of 301 evaluable test results, or 56.5%, were censored at one of the assay reporting limits. Because these values did not represent exact concentrations, arithmetic means and standard deviations of anti-HBs concentration were not calculated. The analysis instead used censoring-aware concentration categories and the prespecified binary seroprotection threshold.

### Participant characteristics according to seroprotection status

Overall, 148 of 296 participants had anti-HBs concentrations ≥10 mIU/mL, corresponding to a seroprotection prevalence of 50.0% (95% confidence interval [CI] 44.3–55.7%). Seroprotection was observed in 79 of 163 females, giving a prevalence of 48.5% (95% CI 40.9–56.1%), and in 69 of 133 males, giving a prevalence of 51.9% (95% CI 43.5–60.2%).

The lowest seroprotection prevalence was observed among participants aged 18–29 years, at 35.3% (95% CI 25.0–47.2%). Prevalence increased across the 30–39, 40–49, and 50–59-year age groups, reaching 71.1% (95% CI 56.6–82.3%) among participants aged 50–59 years. Seroprotection prevalence was 60.0% (95% CI 40.7–76.6%) among those aged ≥60 years. The estimate for participants younger than 18 years was based on only eight observations and was correspondingly imprecise **(Table 1 and Fig 2).**

**Fig 2.**
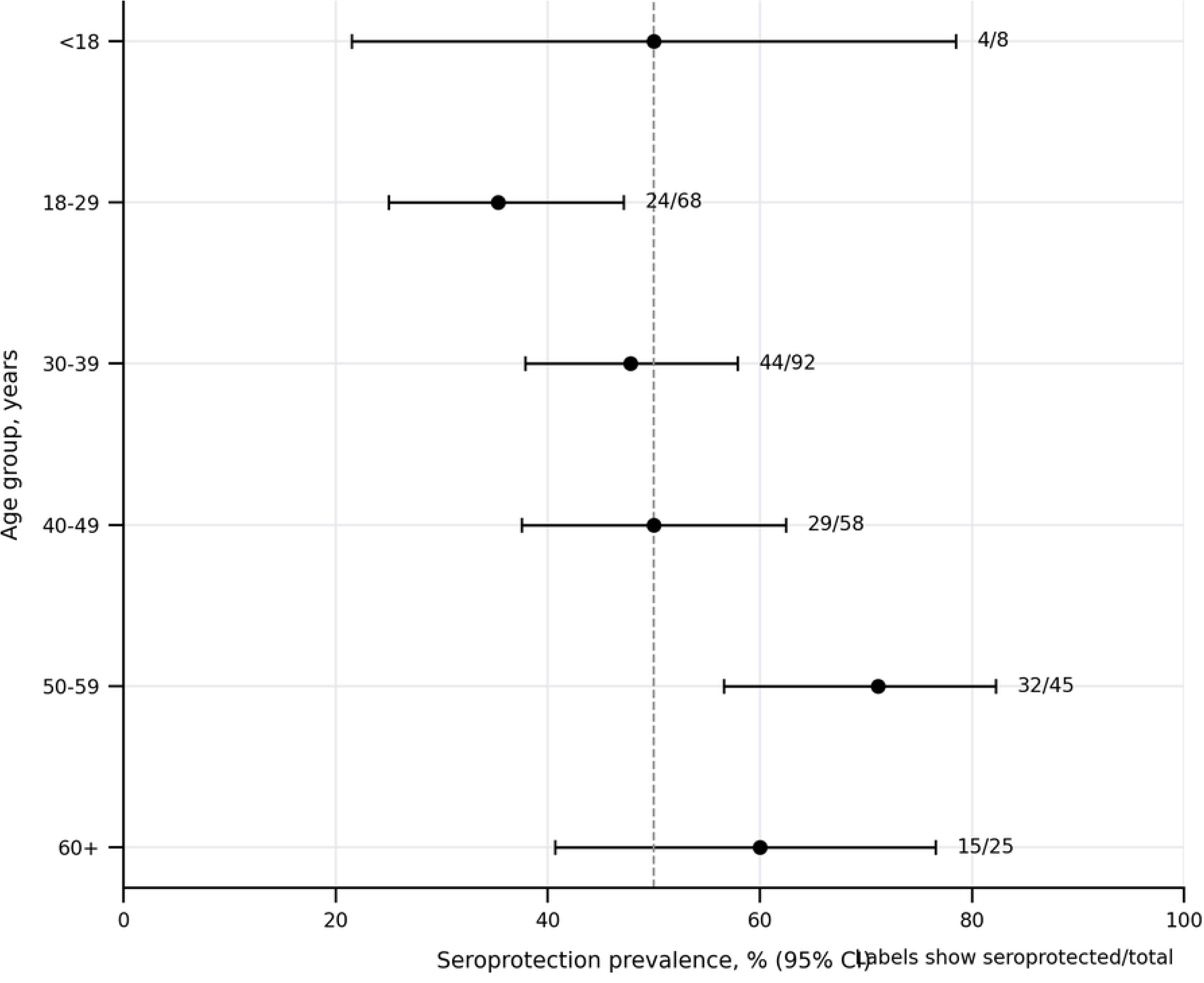
Prevalence of anti-HBs seroprotection by age group. Points indicate the percentage of participants with anti-HBs concentrations at least 10 mIU/mL; horizontal bars represent Wilson 95% confidence intervals. Fractions show the number seroprotected divided by the total number in each age group. The dashed vertical line denotes the overall prevalence of 50.0%. Anti-HBs, antibody to hepatitis B surface antigen; CI, confidence interval.

**Table 1.** Participant characteristics according to anti-HBs seroprotection status.

| Characteristic | Overall,<br>N=296 | Anti-HBs <10<br>mIU/mL, n=148 | Anti-HBs ≥10<br>mIU/mL, n=148 | Seroprotection<br>prevalence, % (95%<br>CI) |
| --- | --- | --- | --- | --- |
| **Age, years** |  |  |  |  |
| **Median (IQR)** | 36 (29–49) | 34 (27–44) | 41 (32–51) | Not applicable |
| **Mean (SD)** | 38.9 (13.4) | 36.4 (12.7) | 41.4 (13.7) | Not applicable |
| **Range** | 14–81 | 15–79 | 14–81 | Not applicable |
| **Sex, n (%)** |  |  |  |  |
| **Female** | 163 (55.1) | 84 (56.8) | 79 (53.4) | 48.5 (40.9–56.1) |
| **Male** | 133 (44.9) | 64 (43.2) | 69 (46.6) | 51.9 (43.5–60.2) |
| **Age group, years, n (%)** |  |  |  |  |
| **<18** | 8 (2.7) | 4 (2.7) | 4 (2.7) | 50.0 (21.5–78.5) |
| **18–29** | 68 (23.0) | 44 (29.7) | 24 (16.2) | 35.3 (25.0–47.2) |
| **30–39** | 92 (31.1) | 48 (32.4) | 44 (29.7) | 47.8 (37.9–57.9) |
| **40–49** | 58 (19.6) | 29 (19.6) | 29 (19.6) | 50.0 (37.5–62.5) |
| **50–59** | 45 (15.2) | 13 (8.8) | 32 (21.6) | 71.1 (56.6–82.3) |
| **≥60** | 25 (8.4) | 10 (6.8) | 15 (10.1) | 60.0 (40.7–76.6) |
| **Calendar quarter, n (%)** |  |  |  |  |
| **First quarter** | 75 (25.3) | 42 (28.4) | 33 (22.3) | 44.0 (33.3–55.3) |
| **Second quarter** | 122 (41.2) | 53 (35.8) | 69 (46.6) | 56.6 (47.7–65.0) |
| **Third quarter** | 77 (26.0) | 43 (29.1) | 34 (23.0) | 44.2 (33.6–55.3) |
| **Fourth quarter** | 22 (7.4) | 10 (6.8) | 12 (8.1) | 54.5 (34.7–73.1) |

Values are n (%) unless otherwise indicated. Percentages in the overall column use all 296 participants as the denominator. Percentages in the anti-HBs <10 mIU/mL and anti-HBs ≥10 mIU/mL columns are column percentages calculated within the respective outcome groups. Seroprotection prevalence is the row-specific proportion with anti-HBs ≥10 mIU/mL; 95% confidence intervals were calculated using the Wilson method. Percentages may not total 100 because of rounding. Anti-HBs, antibody to hepatitis B surface antigen; CI, confidence interval; IQR, interquartile range; SD, standard deviation.

### Factors associated with anti-HBs seroprotection

In unadjusted modified Poisson regression, each 10-year increase in age was associated with a 14% higher prevalence of seroprotection (crude prevalence ratio [PR] 1.14, 95% CI 1.06–1.24; p<0.001). Female participants had a slightly lower estimated prevalence of seroprotection than male participants, although the confidence interval included the null value and was compatible with both a modest reduction and a modest increase in prevalence (PR 0.93, 95% CI 0.74–1.17; p=0.558).

After simultaneous adjustment for age and sex, each 10-year increase in age remained associated with a 14% higher prevalence of seroprotection (adjusted prevalence ratio [aPR] 1.14, 95% CI 1.06–1.24; p=0.001). Sex was not independently associated with seroprotection after adjustment for age (female versus male: aPR 0.97, 95% CI 0.78–1.22; p=0.797) **(Table 2).**

**Table 2.** Crude and adjusted prevalence ratios for factors associated with anti-HBs seroprotection.

| *Factor* | *Crude PR* | *95% CI* | *p-value* | *Adjusted PR* | *95% CI* | *p-value* |
| --- | --- | --- | --- | --- | --- | --- |
| ***Sex*** |  |  |  |  |  |  |
| ***Male*** | *1.00* | *Reference* |  | *1.00* | *Reference* |  |
| ***Female*** | *0.93* | *0.74–1.17* | *0.558* | *0.97* | *0.78–1.22* | *0.797* |
| ***Age, per 10-year increase*** | *1.14* | *1.06–1.24* | *<0.001* | *1.14* | *1.06–1.24* | *0.001* |

*The outcome was anti-HBs seroprotection, defined as an anti-HBs concentration $\geq 10$ mIU/mL.*
*Analyses included 296 participants with an evaluable first anti-HBs test result. Prevalence ratios and 95% confidence intervals were estimated using modified Poisson regression with robust standard errors. The adjusted model included age and sex simultaneously. Age was modelled continuously and rescaled so that the prevalence ratio represents a 10-year increase. Male sex was the reference category. No automated or statistical-significance-based variable-selection procedure was used. Anti-HBs, antibody to hepatitis B surface antigen; CI, confidence interval; PR, prevalence ratio.*

The assumption that age had an approximately linear association with the log prevalence of seroprotection was assessed by adding a quadratic age term to the primary model. There was no evidence that the quadratic term improved model specification (p=0.634), and the linear model had a lower Akaike information criterion than the quadratic model. Age was therefore retained as a continuous linear term expressed per 10-year increase.

### Sensitivity analyses

Adding calendar quarter to the primary model did not materially alter the estimates for age or sex. Each 10-year increase in age remained associated with a higher prevalence of seroprotection (aPR 1.15, 95% CI 1.06–1.24; p<0.001), while the estimate for female versus male participants remained close to the null (aPR 0.98, 95% CI 0.79–1.23; p=0.886).

After adjustment for age and sex, none of the calendar-quarter estimates differed clearly from the first quarter. Compared with the first quarter, the adjusted prevalence ratio was 1.27 for the second quarter (95% CI 0.95–1.70; p=0.107), 0.97 for the third quarter (95% CI 0.68–1.37; p=0.849), and 1.29 for the fourth quarter (95% CI 0.82–2.03; p=0.277). Because we reconstructed five testing dates from accession-number chronology, we treated the calendar-quarter estimates as supportive sensitivity analyses. We did not use them to determine the principal study conclusions.

Restricting the analysis to the 288 participants aged ≥18 years yielded results consistent with the primary analysis. Each 10-year increase in age was associated with a 16% higher adjusted prevalence of seroprotection (aPR 1.16, 95% CI 1.07–1.25; p<0.001), while sex remained unassociated with seroprotection (female versus male: aPR 0.98, 95% CI 0.78–1.23; p=0.835).

When age was modelled categorically, with participants aged 18–29 years as the reference group, seroprotection prevalence was approximately twice as high among participants aged 50–59 years (aPR 2.01, 95% CI 1.38–2.91; p<0.001) and 69% higher among those aged ≥60 years (aPR 1.69, 95% CI 1.07–2.66; p=0.025). Estimates for participants aged 30–39 years, 40–49 years, and <18 years were directionally positive but imprecise. These sensitivity analyses supported the robustness of the primary age association, although the estimates should not be interpreted causally **(Table 3 and Fig 3).**

**Fig 3.**
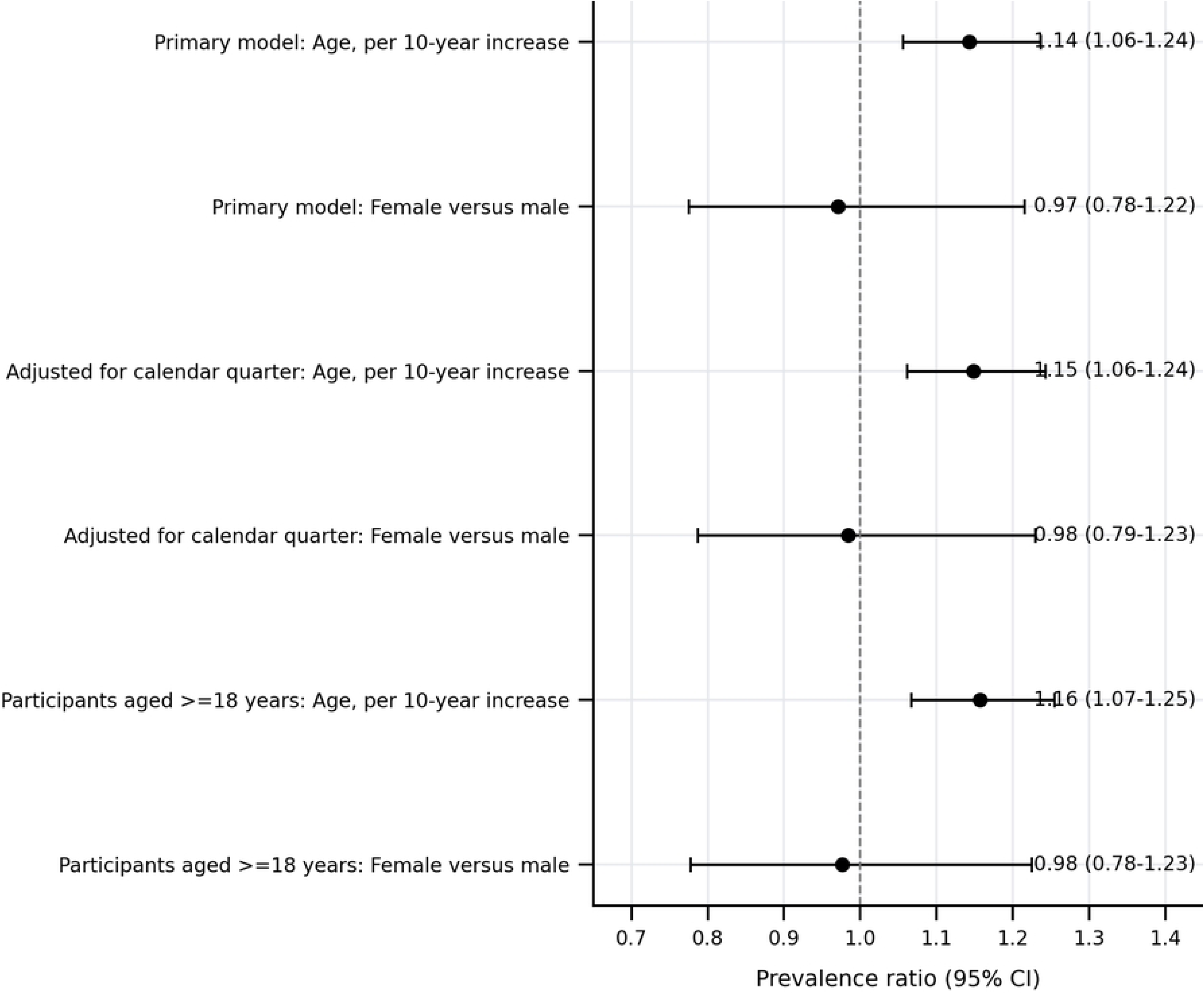
Adjusted prevalence ratios for anti-HBs seroprotection in the primary and sensitivity analyses. Points show adjusted prevalence ratios, and horizontal bars show 95% confidence intervals estimated using modified Poisson regression with robust standard errors. The primary model included age and sex. The calendar-quarter sensitivity model additionally included calendar quarter, and the adult-only model was restricted to participants aged at least 18 years. Male sex was the reference category. The dashed vertical line denotes the null value of 1. Anti-HBs, antibody to hepatitis B surface antigen; CI, confidence interval.

**Table 3.** Sensitivity analyses of factors associated with anti-HBs seroprotection.

| Sensitivity analysis | Factor | Adjusted PR | 95% CI | p-value |
| --- | --- | --- | --- | --- |
| **Additional adjustment for calendar quarter** | Female versus male | 0.98 | 0.79–1.23 | 0.886 |
|  | Age, per 10-year increase | 1.15 | 1.06–1.24 | <0.001 |
|  | Second versus first quarter | 1.27 | 0.95–1.70 | 0.107 |
|  | Third versus first quarter | 0.97 | 0.68–1.37 | 0.849 |
|  | Fourth versus first quarter | 1.29 | 0.82–2.03 | 0.277 |
| **Restricted to participants aged ≥18 years** | Female versus male | 0.98 | 0.78–1.23 | 0.835 |
|  | Age, per 10-year increase | 1.16 | 1.07–1.25 | <0.001 |
| **Age modelled categorically** | 30–39 versus 18–29 years | 1.35 | 0.91–1.99 | 0.133 |
|  | 40–49 versus 18–29 years | 1.41 | 0.93–2.13 | 0.103 |
|  | 50–59 versus 18–29 years | 2.01 | 1.38–2.91 | <0.001 |
|  | ≥60 versus 18–29 years | 1.69 | 1.07–2.66 | 0.025 |
|  | <18 versus 18–29 years | 1.43 | 0.67–<br>3.06 | 0.357 |
|  | Female versus male | 0.96 | 0.77–<br>1.20 | 0.729 |

*All estimates were obtained using modified Poisson regression with robust standard errors. The* *calendar-quarter model included continuous age, sex, and calendar quarter. The adult-only model* *included continuous age and sex. The categorical-age model included age group and sex. Male sex,* *the first calendar quarter, and age 18–29 years were the respective reference categories. Anti-HBs,* *antibody to hepatitis B surface antigen; CI, confidence interval; PR, prevalence ratio.*

## Discussion

### Principal findings

In this retrospective laboratory-based cross-sectional study, 50.0% of healthcare workers undergoing anti-HBs testing at Eastgate Clinic had antibody concentrations at or above the operational seroprotection threshold of 10 mIU/mL. Conversely, half had concentrations below this threshold, including 44.6% whose results were below the assay’s lower reporting limit of 2 mIU/mL. Seroprotection prevalence did not differ materially by sex. In contrast, age was positively associated with seroprotection: each 10-year increase in age was associated with a 14% higher adjusted prevalence. This association remained consistent after adjustment for calendar quarter, restriction of the analysis to adults, and alternative modelling of age as a categorical variable.

These findings identify a substantial burden of non-protective anti-HBs results within the population accessing testing at Eastgate Clinic. They do not, however, indicate that half of all healthcare workers in Windhoek are unprotected or inadequately vaccinated. The study population was defined by receipt of laboratory testing rather than probability-based sampling of the healthcare workforce. The prevalence estimate therefore characterises healthcare workers who entered this testing pathway during 2025 and should not be extrapolated directly to untested healthcare workers in Windhoek or Namibia.

### Interpretation in relation to existing evidence

Healthcare workers are at occupational risk of HBV exposure through percutaneous injuries, mucosal contact, and exposure of non-intact skin to blood or other potentially infectious body fluids. Across African settings, studies have documented persistent HBV infection risks, incomplete vaccination, and uneven access to occupational-health services, post-vaccination testing, and post-exposure management [3–6]. In Namibia, previous research found that healthcare workers were generally aware of hepatitis B, but that implementation and use of national prevention and control guidance remained insufficient [7]. The present findings extend this evidence by demonstrating that a substantial proportion of healthcare workers reaching an anti-HBs testing service had antibody concentrations below the conventional threshold used to define an adequate post-vaccination serological response.

Direct comparison with previous studies requires caution. Published studies among healthcare workers have differed in their populations, vaccination histories, timing of antibody testing, assay platforms, and definitions of seroprotection. Some studies were undertaken specifically among workers with documented completion of a hepatitis B vaccine series, whereas the present laboratory register did not include vaccination status or test indication. The 50.0% seroprotection prevalence observed here therefore cannot be interpreted as a vaccine-response rate. It may reflect a heterogeneous combination of unvaccinated workers, incomplete vaccination, testing before vaccination, testing after an appropriately timed vaccine series, delayed testing after vaccination, resolved natural infection, booster responses, and true primary vaccine non-response.

The higher prevalence of seroprotection at older ages warrants particularly careful interpretation. Measurable anti-HBs concentrations may decline after vaccination, meaning that older age would not necessarily be expected to correspond to higher titres in a uniformly vaccinated population tested at comparable intervals after vaccination [1,14]. The observed age association is more plausibly explained by differences in the composition and testing histories of the age groups. Older healthcare workers may have had more opportunities to receive occupational vaccination or booster doses, may have been preferentially referred for testing after documented vaccination, or may have acquired anti-HBs following resolved natural infection. Conversely, younger participants may have included students, trainees, or recent entrants to the health workforce who were evaluated before completing a vaccination series.

The categorical age analysis supported the overall pattern but also indicated that the relationship should not be interpreted as an uninterrupted biological increase across the life course. Compared with participants aged 18–29 years, the adjusted prevalence was approximately twice as high among those aged 50–59 years and 69% higher among those aged 60 years or older. Estimates for the intermediate age groups were directionally positive but imprecise. The absence of evidence for departure from linearity in the continuous-age model supported the use of age per 10-year increase as the parsimonious primary specification. Nevertheless, the association remains descriptive and may reflect selection, vaccination history, cumulative exposure, testing indication, or other unmeasured factors rather than an independent effect of ageing.

Seroprotection prevalence was similar among female and male participants. The adjusted estimate was close to the null, and its confidence interval was compatible with both a modest reduction and a modest increase in prevalence among females. These findings provide no compelling evidence for sex-specific differences in the tested population. Occupational-health responses should therefore prioritise documented vaccination history, exposure risk, vaccine-series completion, appropriate test timing, and follow-up rather than sex alone.

Calendar-quarter estimates also did not provide evidence of a consistent temporal pattern. Although seroprotection prevalence appeared higher in the second and fourth quarters than in the first quarter, the corresponding confidence intervals were wide and included the null value after adjustment for age and sex. These differences may reflect variation in testing volume, referral practices, workplace screening activities, or the composition of workers tested during different periods. Calendar quarter should therefore be regarded as a service-delivery characteristic rather than a biological determinant of seroprotection.

### Interpretation of the anti-HBs threshold

The 10 mIU/mL threshold has its clearest clinical interpretation when anti-HBs is measured using a validated quantitative assay 1–2 months after completion of a documented hepatitis B vaccination series [1,14]. Under these conditions, a concentration of at least 10 mIU/mL is considered evidence of an adequate vaccine-induced serological response. Healthcare workers with reasonably anticipated exposure to blood or body fluids should have vaccination and, where indicated, post-vaccination testing documented [14–16].

Interpretation is less straightforward when the vaccination series and timing of testing are unknown. Among immunocompetent individuals who previously achieved an adequate post-vaccination response, antibody concentrations may subsequently decline below 10 mIU/mL while immune memory persists [14]. A result below 10 mIU/mL obtained years after vaccination therefore does not, by itself, establish primary vaccine failure or complete susceptibility. Conversely, anti-HBs positivity alone does not establish vaccine-derived immunity because anti-HBs may also follow resolved natural infection. HBsAg and total anti-HBc are needed to distinguish vaccine-associated immunity, past infection, possible current infection, and susceptibility more comprehensively [8,9,15].

Accordingly, the term seroprotection is used operationally in this study to denote an anti-HBs concentration ≥10 mIU/mL. It should not be interpreted as a complete clinical classification of HBV immunity. Similarly, a concentration below 10 mIU/mL should be described as a non-protective anti-HBs result rather than definitive evidence of vaccine non-response.

### Occupational-health and policy implications

The findings support strengthening the hepatitis B occupational-health pathway for healthcare workers. Health facilities should establish systems that document hepatitis B vaccination status at entry into employment, professional training, or clinical placement; identify workers with occupational exposure to blood or body fluids; provide vaccination without avoidable financial or administrative barriers; and document post-vaccination serological testing where indicated [2,14–16]. Vaccination of healthcare workers is not only an individual protective measure but also an element of infection prevention and control, patient safety, and health-system resilience. The World Health Organization recommends integrating healthcare-worker vaccination into occupational-health and infection-prevention programmes and providing recommended vaccines without cost to healthcare workers [2].

A non-protective anti-HBs result should trigger a structured review rather than automatic classification as vaccine failure. This review should determine whether the worker has written, dated evidence of vaccination; whether the primary series was completed; whether testing occurred at an appropriate interval after the final dose; whether additional vaccination is indicated; and whether post-exposure management is required. Guidance for healthcare personnel emphasises written documentation of vaccine uptake and post-vaccination serological response among workers with reasonably anticipated occupational exposure [14–16].

Laboratory request systems should be revised to capture, at minimum, healthcare-worker cadre, employing facility, reason for testing, vaccination-series status, dates of vaccine doses, and whether testing is pre-vaccination, post-series, post-booster, or post-exposure. These variables would permit clinically meaningful interpretation of anti-HBs results and support surveillance of vaccination completion and follow-up. Where clinically indicated, the use of the HBV triple panel comprising HBsAg, anti-HBs, and total anti-HBc would allow more complete classification of infection and immunity status [8,9,15].

The findings also indicate an opportunity to improve the linkage between laboratory testing and occupational-health action. A laboratory result should not represent the endpoint of the pathway. Systems are needed to confirm that results are communicated, vaccination or additional doses are offered where indicated, post-exposure prophylaxis is initiated when required, and repeat testing is completed within the recommended interval. Such a system would transform isolated laboratory measurements into an actionable occupational-health programme.

### Strengths and limitations

This study has several strengths. It analysed routine anti-HBs testing over a full calendar period and retained the original reporting structure of the laboratory results. Values below and above the assay range were treated as censored rather than as exact concentrations, avoiding biased calculation of arithmetic means and standard deviations. The primary analysis was conducted at the individual level, with the first chronological test retained for participants with repeat testing. This preserved the independence of observations and avoided giving greater analytical weight to individuals tested more than once.

The statistical strategy was aligned with the outcome distribution. Because seroprotection occurred in half of participants, modified Poisson regression with robust standard errors was used to estimate prevalence ratios rather than odds ratios. Age was retained as a continuous variable in the primary model, thereby preserving information and limiting unnecessary parameters. Its functional form was assessed, and there was no evidence that adding a quadratic term improved model specification. The principal age association was also consistent after adjustment for calendar quarter, restriction to adults, and categorical modelling of age. Data-cleaning decisions were documented, missing outcomes were not imputed, and confidence intervals were presented for prevalence and association measures.

These strengths should be considered alongside several limitations. First, the study included healthcare workers who underwent testing at one private healthcare facility. Referral for anti-HBs testing was unlikely to be random. Participants may have been tested because of employment requirements, vaccination, clinical training, occupational exposure, a previous non-protective result, or individual concern. Selection into testing could therefore have produced a study population that differed materially from the wider healthcare workforce. The findings should not be interpreted as estimating workforce-wide seroprotection prevalence in Windhoek or Namibia.

Second, healthcare-worker status was not directly encoded as a variable in the supplied analytical spreadsheet. Eligibility must be confirmed against request forms, occupational-health records, referral documentation, or another reliable source before submission. Misclassification of laboratory clients as healthcare workers would threaten the validity of the study population definition.

Third, the completeness of record capture could not be established from the spreadsheet alone. The register should be reconciled with the laboratory information system, analyser logs, billing records, or other source documentation to confirm that all eligible anti-HBs tests performed during 2025 were included. If some tests were omitted systematically, the prevalence estimates may be affected by selection bias.

Fourth, the available data contained only age, sex, test date, and anti-HBs result. Occupational cadre, employing facility, patient-contact status, exposure history, vaccination status, number and timing of vaccine doses, testing indication, HIV status, immunosuppressive conditions, HBsAg, total anti-HBc, and clinical follow-up were unavailable. The multivariable analysis could therefore address only recorded demographic and temporal factors. Residual confounding is likely, and the age association should not be interpreted as causal.

Fifth, anti-HBs alone cannot distinguish vaccine-induced immunity from resolved natural infection. In addition, a concentration below 10 mIU/mL cannot be equated unconditionally with susceptibility or primary vaccine non-response when previous vaccine response and timing of testing are unknown. The study consequently estimated an operational seroprotection prevalence rather than vaccine effectiveness, vaccination coverage, or the prevalence of definitive HBV susceptibility.

Sixth, repeat tests were linked using normalised names and supporting demographic and chronological information rather than a stable unique patient identifier. This approach may have failed to identify some repeat individuals whose names were recorded differently, while people with identical or highly similar names could potentially have been linked incorrectly. One apparent repeat pair had discordant recorded sex, underscoring the limitations of deterministic linkage using routine identifiers.

Seventh, five test dates were inconsistent with the study period or the chronological sequence of laboratory accession numbers and were provisionally reconstructed using adjacent laboratory records. The affected observations were flagged for verification against the original source register. This uncertainty did not affect classification of the primary seroprotection outcome or the principal age-and-sex regression model, but it may have affected assignment of participants to calendar quarters. The quarter-adjusted analysis should therefore be regarded as a sensitivity analysis rather than a definitive assessment of temporal variation.

Finally, some subgroup estimates were based on small numbers. This was particularly evident among participants younger than 18 years and those tested during the fourth quarter, for whom confidence intervals were wide. These estimates should be interpreted as descriptive. The study was not designed or powered to detect effect modification, and no subgroup interaction findings are advanced as principal conclusions.

## Conclusion

Among healthcare workers undergoing anti-HBs testing at Eastgate Clinic during 2025, half had antibody concentrations ≥10 mIU/mL and half had concentrations below this operational seroprotection threshold. Older age was associated with a higher prevalence of seroprotection, whereas sex was not independently associated with the outcome. These findings identify an important gap within the occupational-health testing pathway but do not establish workforce-wide immunity prevalence, vaccination coverage, or vaccine effectiveness.

The findings support systematic verification of hepatitis B vaccination, completion of recommended vaccine schedules, appropriately timed post-vaccination testing, and documented follow-up of healthcare workers with non-protective results. Linking future laboratory data to occupational characteristics, vaccination histories, testing indications, complete HBV serological profiles, and subsequent clinical actions would permit more precise classification of immunity and susceptibility and provide a stronger empirical basis for occupational-health policy in Namibia.

## Data Availability Statement

The de-identified minimal dataset underlying the findings reported in this article, the accompanying data dictionary, and the source values used to generate the figures will be available from the designated public repository upon publication. Direct identifiers, laboratory accession numbers, the restricted linkage file, and exact test dates that could increase the risk of participant re-identification are not included in the deposited dataset.

## Author contributions

**Conceptualization:** Talent Farai Mafumhe, Hamufare Dumisani Mugauri.

**Data curation:** Talent Farai Mafumhe, Hamufare Dumisani Mugauri.

**Formal analysis:** Hamufare Dumisani Mugauri.

**Investigation:** Talent Farai Mafumhe.

**Methodology:** Talent Farai Mafumhe, Vonai Charamba, Onyemaechi Okpara Azu, Hamufare Dumisani Mugauri.

**Project administration:** Talent Farai Mafumhe.

**Resources:** Talent Farai Mafumhe.

**Supervision:** Onyemaechi Okpara Azu, Hamufare Dumisani Mugauri.

**Validation:** Talent Farai Mafumhe, Vonai Charamba, Onyemaechi Okpara Azu, Hamufare Dumisani Mugauri.

**Visualization:** Hamufare Dumisani Mugauri.

**Writing, original draft:** Talent Farai Mafumhe, Hamufare Dumisani Mugauri.

**Writing, review and editing:** Talent Farai Mafumhe, Vonai Charamba, Onyemaechi Okpara Azu, Hamufare Dumisani Mugauri.

## Acknowledgments

The authors acknowledge the laboratory and administrative staff of Eastgate Clinic for maintaining the routine records used in this study.

